# Variation in colorectal cancer treatment and outcomes in Scotland: real world evidence from national linked administrative data

**DOI:** 10.1101/2022.01.28.22270027

**Authors:** E Lemmon, C Hanna, K Diernberger, PS Hall

## Abstract

**Background:** Colorectal cancer (CRC) is the third most common type of cancer in Scotland and the second leading cause of cancer death. Despite improvements in CRC survival over time, Scotland lags behind its UK and European counterparts. Linked administrative datasets can provide a real-world representation of current care as a basis for evaluation of new interventions or policies and to understand variation in care and outcomes. In this study, we aim to provide up to date, population level evidence on CRC treatment and survival in Scotland for patients treated with curative or palliative intent as a basis for the understanding of variation, and to provide data to underpin the evaluation of new treatments.

**Methods:** We conducted a retrospective analysis of adults with an incident CRC registered on the Scottish Cancer Registry (ICD-10 codes C18-20) between January 2006 and December 2018. Data on patients with incident CRC was linked to hospital inpatient records allowing description of their demographic, diagnostic and treatment characteristics. For a curative cohort (n = 26,204) Cox-Proportional Hazards regression models were used to assess the factors affecting overall survival (OS) and CRC specific survival (CRCS).

**Results:** Overall, 32,691 (73%) and 12,184 (27%) patients had a diagnosis of colon and rectal cancer respectively. On average, patients with colorectal cancer had two hospital inpatient stays in the five-years pre-diagnosis. Chemotherapy was used in 42% with colon cancer and 30% with colon cancer. Radiotherapy use was 2% and 39% respectively. Five year OS (CRCS) within the curative cohort was 71% (81%) and 75% (82%) for patients with colon and rectal cancer respectively. After accounting for patient and tumour characteristics, survival outcomes demonstrated significant geographical variation.

**Conclusions:** National linked administrative datasets have the ability to provide real-world representation of the treatments and outcomes for patients with cancer. In a Scottish population of curative patients with CRC, there was significant variation in survival depending on sex and geography.

## 1. INTRODUCTION

Colorectal cancer (CRC) is the third most common type of cancer for men and women in Scotland and the second leading cause of cancer death (Public Health Scotland, 2020; Information Services Division Scotland, 2019). It is projected that the number of new CRC cases in Scotland will increase by 43% by 2023-27 compared to 2008-12(Information Services Division Scotland, 2015).

Despite improvements in CRC survival over time, Scotland lags behind its UK and European counterparts (De Angelis et al., 2014; Ferlay et al., 2018). Previous research has shown that socio-economic deprivation and remoteness factors, for example distance from a cancer centre, are significantly associated with poorer survival outcomes in Scotland (Shack et al., 2007; Campbell et al., 2000) and in the UK (Lyratzopoulos et al., 2011; Smith et al., 2006; Møller et al., 2012). However, there are considerable differences in conclusions across studies and the mechanisms behind the observed relationships remain unclear (Paterson et al., 2014; Brewster et al., 2001; Hole and McArdle, 2002). Possible explanations for the differences in survival outcomes observed across geographies and patient cohorts include delayed presentation, stage at presentation, treatment and co-morbidity.

Research in this area, and ultimately patient outcomes, may be improved by utilising the vast amounts of administrative healthcare data that are collected routinely as part of the delivery of patient care (Lemmon et al., 2021). It provides an opportunity to generate evidence with a high degree of external validity, being entirely representative of current care (Connelly et al., 2016). Administrative records, unlike trial or observational research data, provide detailed evidence on whole populations, over extended periods of time. This enables research that is inclusive of patient groups who are traditionally harder to reach, while exploiting the longitudinal nature of the data offers insight into changes that occur over time.

As well as real-world representativeness, a greater breadth of information can be obtained by linkage between datasets. Previous research has shown that administrative data provide a sufficiently accurate source of information to describe patients with cancer and their outcomes (Goldsbury et al., 2012). At the same time, linkage between datasets can provide a richer characterisation of patient needs, which is essential to understand the mechanisms underlying differences in treatment and outcomes.

In England, linked administrative datasets have been used within the CRC context to investigate routes to diagnosis (Pearson et al., 2019); explore provider differences in post-colonoscopy recurrence rates (Burr et al., 2019); explain variation in treatment and outcomes (Morris et al., 2010; Taylor et al., 2021); describe management of disease (Birch et al., 2019) and much more. In Scotland, there are very few CRC studies using linked administrative datasets to investigate CRC treatment and many have used administrative data from a single geographic area e.g. (Paterson et al., 2014; Robertson et al., 2004; Hole and McArdle, 2002). Further, published, population level statistics on survival tend to group stages and/or disease sites together, rather than look at outcomes for patients who are treated on a curative pathway.

In this study, we contribute to existing research by providing up to date, population level evidence on CRC treatment and survival outcomes for patients who are diagnosed with CRC in Scotland. We use a newly established, unique CRC dataset, which links demographic data to the Scottish Cancer Registry and routine hospital admissions data. Full details of this dataset are described elsewhere (Hanna et al., 2021). In what follows, we firstly describe the demographic, diagnostic and treatment characteristics of patients with colon and rectal cancer. Secondly, for those patients treated with curative intent, we estimate their survival and assess the factors affecting their overall survival and CRC specific survival.

## 2. METHODS

The study population consisted of adults having an incident CRC registered on the Scottish Cancer Registry (ICD-10 codes C18, C19 and 20) between January 2006 and December 2018. Approval for the study was granted by the Public Benefit and Privacy Panel (PBPP) for health and social care, project number 1718-0026. The study meets the requirements for ethical approval set out by the East of Scotland NHS Research Ethics Service for the analysis of secondary National Services of Scotland (NSS) data.

### 2.1. Data

We used the Scottish Cancer Registry (Scottish Morbidity Record 06 (SMR06)) to identify a cohort of patients diagnosed with CRC. National Records of Scotland (NRS) deaths data was used to provide survival outcomes and inpatient and day case hospital admissions data (SMR01) provided information on comorbidities and hospital use prior to cancer diagnosis. These three datasets were linked via a pseudonomysied patient identifier.

#### Scottish Cancer Registry (SMR06)

The Scottish Cancer Registry dataset includes information on all new diagnoses of cancer occurring within Scotland. This data is collected by Public Health Scotland (PHS) and contains diagnostic, staging and treatment information. Each SMR06 record for a patient corresponds to a unique cancer diagnosis for that individual.

In this study, we had access to all SMR06 records for patients who had a diagnosis of CRC (International Disease Classification 10th Revision (ICD-10) codes C18, C19 or C20) between January 2006 and December 2018.

#### Inpatient and day case admissions (SMR01)

The SMR01 dataset contains episode level data for all general/acute inpatient or day cases in Scottish NHS hospitals or Scottish NHS beds in non-NHS-institutions. This study used all patient SMR01 records between 1997 and 2018, for any patient present in the study SMR06 dataset.

#### National Records of Scotland (Deaths)

The NRS are responsible for the registration of all life events occurring in Scotland including births, deaths, marriages, civil partnerships and adoptions. For the purposes of this study, NRS vital events data on births and deaths was used to obtain patient date of birth, sex, date and cause of death.

### 2.2. Cohort derivation

The retrospective cohort was derived from the SMR06 database. Prior to matching patient records to their SMR01 and deaths records, a number of exclusion criteria were applied. These criteria are described in Fig. 1. In particular, patients were excluded if their diagnosis of CRC occurred before the study period using the flag for an historic CRC diagnosis (1.3% of records). Next, we excluded all non-CRC diagnoses (19.6% of records) and any CRC diagnosis that was diagnosed at autopsy (1.4% of records). Following this, using the incidence date, all secondary CRC diagnoses were excluded (1% of records). Further, patients were excluded in the event that they had more than one CRC diagnosis (i.e. different ICD-10 codes) with the same incidence date (less than 1% of records). In the event that a patient had more than one of the same CRC diagnosis (i.e. the same ICD-10 code) with the same incidence date, staging information was used to exclude the least severe diagnosis and in the event that severity was identical, the records with the completion of most variables were kept (less than 1% of records). Finally, where incidence date, type of CRC, severity and data fullness were identical, duplicate records were randomly dropped.

**Figure 1:**
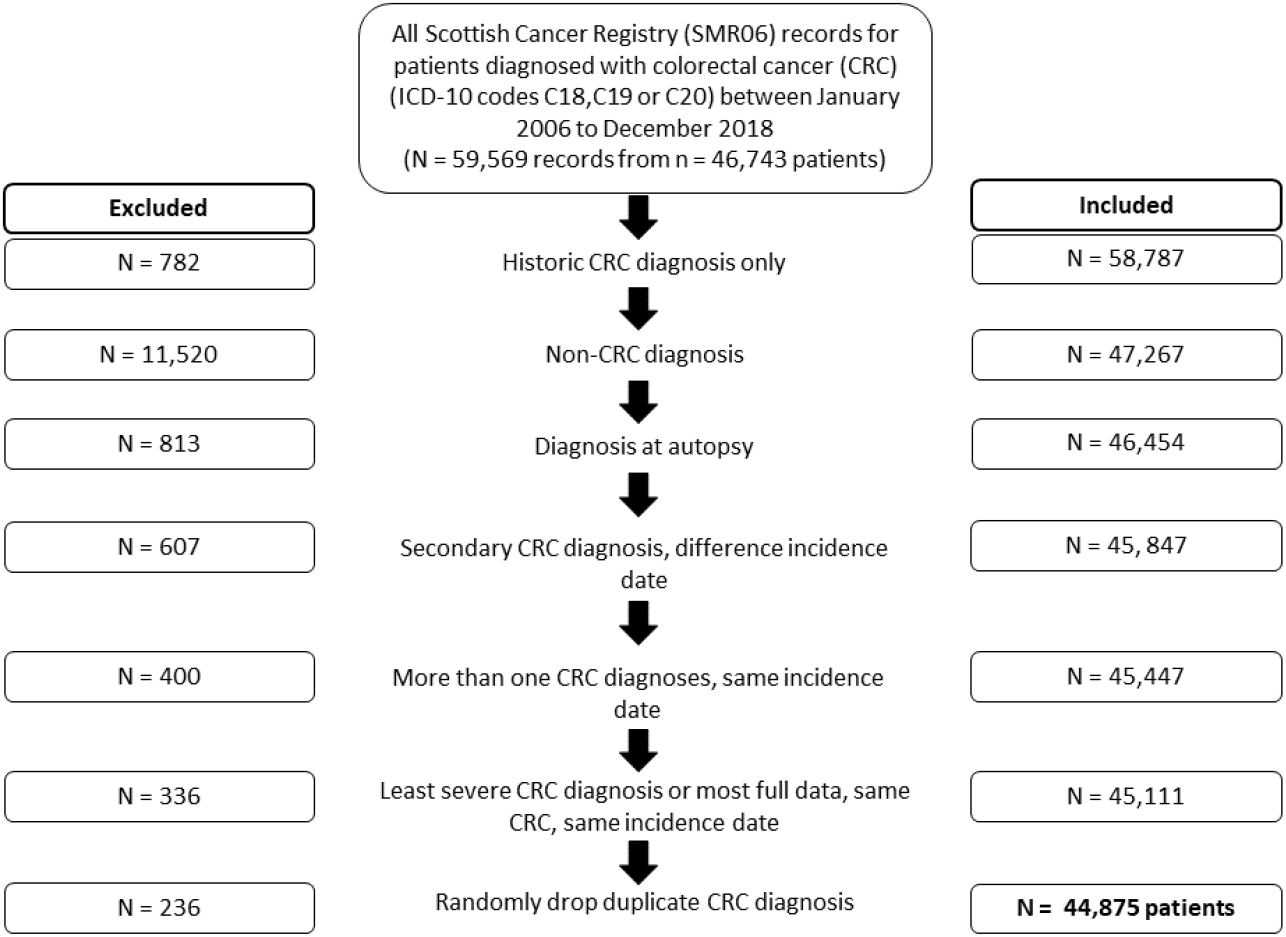
Cohort Derivation Flow Chart

Following exclusions, a total of N = 44,875 patients remained. This cohort was then linked to the SMR01 and deaths records to obtain pre-diagnosis co-morbidity, hospital admissions information and survival outcomes.

### 2.3. Descriptive analysis

The full cohort was characterised by descriptive statistics of their demographics, diagnosis and treatment. The variables included are described in Table 1 below.

**Table 1:** Demographics

| Variable | Description |
| --- | --- |
| **Demographics** |  |
| Sex | Binary sex indicator to indicate if the patient is male or female |
| Age | 10 year age bands (18-34, 35-44, 45-54, 55-64, 65-74, 75-84, 85+) based on age at diagnosis |
| SIMD Quintile | Scottish Index of Multiple Deprivation Quintile from most deprived (1) to least deprived (5) |
| Urban/Rural | Binary indicator to indicate the rurality of the usual residence of the patient. Comes from the Scottish Government's six-fold urban/rural classification. Urban locations are defined as large urban areas, other urban areas or accessible small towns. Remote/rural are defined as remote small towns, accessible rural or remote rural. |
| Cancer Network | An indicator of the Managed Cancer Network (MCN) in which the patient was diagnosed. There are three MCNs in Scotland including South of Scotland Cancer Network (SCAN), West of Scotland Cancer Network (WoSCAN) and the North of Scotland Cancer Network (NoSCAN) |
| **Diagnosis** |  |
| Stage (Duke's stage) | Indicates the extent of spread of the invasive tumour at diagnosis in terms of the pathological and/or clinical findings (Stage I, II, III, IIII or Unknown) |
| Method 1st detection | Categorical indicator to indicate how the tumour was first detected (Screening examination, incidental finding, clinical presentation, interval cancer or other, not known. |
| Year of diagnosis | Three year bands for the year of diagnosis (2006-08, 2009-11, 2012-14 and 2015-18) |
| **Treatment** |  |
| Therapy objectives | Categorical indicator to indicate the treatment intent (curative, palliative or unknown) |
| Chemotherapy | Binary indicator to indicate if the patient has had systemic chemotherapy treatment |
| Radiotherapy | Binary indicator to indicate if the patient was treated with radiotherapy |
| Surgery | Binary indicator to indicate if the patient was treated with surgery |
| Inpatient episodes | Mean number of hospital inpatient episodes in the five years pre-diagnosis (derived from SMR01) |
| Quan morbidity score | Mean morbidity score (Quan et al., 2005) in the five years pre-diagnosis (derived from SMR01) using Charlson indicators with Quan weights (excludes cancer and metastatic cancer). |

Since the disease trajectories and treatment pathways are quite different for patients with rectal cancer (ICD.10 code C20) compared to patients with colon cancer (ICD-10 codes C18 and C19), analysis was carried out separately for these two disease sites.

### 2.4. Statistical analysis

All patients who were treated on a curative pathway were included in the survival analysis. We defined this curative cohort using a number of variables. Firstly, we removed those who did not undergo surgery or had an unknown surgery status (n = 11,506). Next, we removed those who had palliative or unknown therapy objectives (n = 4,618). Following this, we removed all patients who were diagnosed with stage IIII (n = 725) or unknown disease (n = 1,517). Finally, we removed patients where the chemotherapy and radiotherapy treatment variables were missing (n = 305). This resulted in a final curative cohort of 26,204 patients. Analysis of patients with metastatic or incurable CRC will be published separately.

We defined two survival end points: Overall survival (OS) and CRC survival (CRCS). OS was the interval between the date of diagnosis and the date of death, censored at the date of the most recent death observed in the data set in December 2018. CRCS was defined as the interval between the date of diagnosis and the date of death, where CRC was classified as the underlying cause of death. Those lost to follow up were censored in the same way as outlined for OS with the additional censoring of date of death for those who died from non-CRC related causes. Survival curves were estimated using the Kaplan-Meier method.

Finally, we conducted univariable and multivariable analysis to examine the factors associated with OS and CRCS using Cox-Proportional Hazards models. Multivariable models were adjusted for sex, age group, cancer network, SIMD quintile, Duke’s stage, urban/rural, year of diagnosis, mean number of inpatient episodes and Quan comorbidity score in the five years pre-diagnosis.

## 3. RESULTS

### 3.1. Patient characteristics

The final study population included 44,875 patients. Patient characteristics are described in Table 2. Overall, 32,691 (73%) of patients had a diagnosis of colon cancer and 12,184 (27%) had a rectal cancer diagnosis. The majority of patients diagnosed with colon and rectal cancer were aged over 65.

**Table 2:**
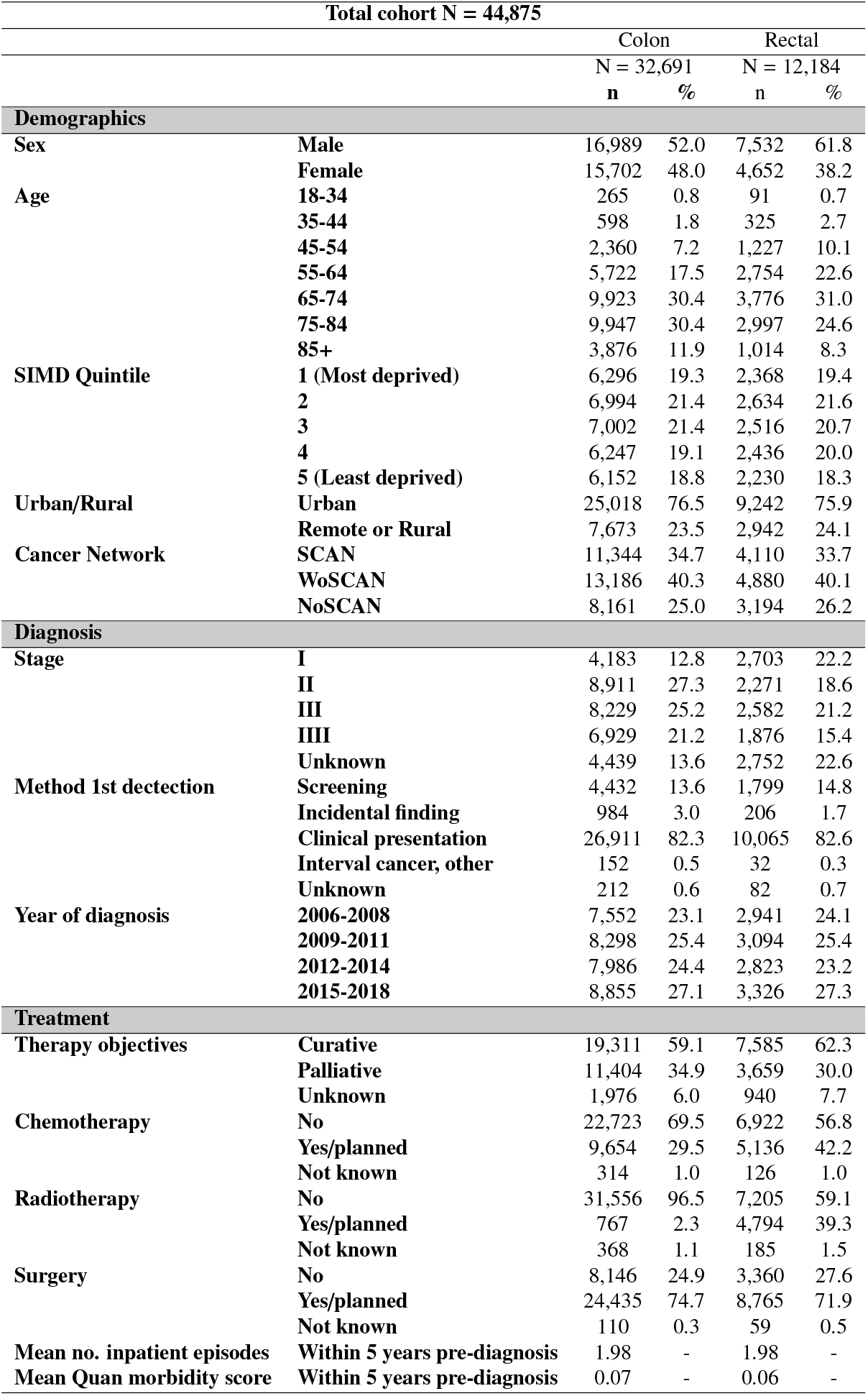
Patient Characteristics

The majority of patients lived in urban areas (around 76% for both cancers). WoSCAN is the largest MCN in Scotland and as expected, it accounts for the largest proportion of CRC patients (40%), followed by SCAN (35% colon and 34% rectal) and NoSCAN (25% colon and 26% rectal).

In terms of the Scottish Index of Multiple Deprivation (SIMD), there is little variation across quintiles though for both cancer groups, a slightly lower proportion of patients come from the least deprived quintile (quintile five) compared to the most deprived quintile (quintile one).

Hospital admissions records from the five years prior to diagnosis show that patients diagnosed with colon cancer and rectal cancer both had an average Quan co-morbidity 0 (Quan et al., 2005).

### 3.2. Diagnosis

The majority of patients were diagnosed via clinical presentation, with approximately 14% of patients with colon cancer and 15% of patients with rectal cancer, diagnosed via screening. There were differences in stage of disease at diagnosis depending on disease site. For patients with rectal cancer, 21% had stage I disease and 19% had stage II disease at diagnosis compared to 13% and 27% respectively for patients with colon cancer. More patients with rectal cancer had an unknown stage of disease recorded at diagnosis (24% versus 15% for colon cancer). Diagnoses were approximately evenly distributed throughout the study period with between 23 and 25% of patients being diagnosed within each three year period and 27% in the final four years.

### 3.3. Treatment

The majority of patients were treated with curative intent according to the therapy objectives variable recorded in the cancer registry. Specifically, 59% of patients diagnosed with colon cancer and 62% of patients with rectal cancer, with a smaller proportion treated with palliative intent (35% and 30% respectively). A small proportion of patients had unknown therapy objectives, 6% and 8% respectively. Accordingly, over 70% of patients with colon and rectal cancer underwent or had planned surgery.

The treatment for patients with colon cancer differs compared to those with rectal cancer. For the whole cohort, patients with a colon cancer diagnosis were less likely to receive chemotherapy (30%) compared to patients with rectal cancer (42%), and they were much less likely to receive radiotherapy (2% versus 39%).

On average, for both cancer types, patients had 11 inpatient or day case episodes in the five years prior to their diagnosis.

### 3.4. Survival

Table 3 presents survival outcomes for the curative cohort (n = 26,204). In total, 9,094 patients died between January 2006 and December 2018. Of these, 36% (6,930) of patients with colon cancer 31% (2,164) of patients with rectal cancer died within the follow-up period. On average, patients were followed up for 1,051 days. There were 5,040 CRC specific deaths. Around 19% of both patients with colon cancer (3,722) and rectal cancer (1,318) died from CRC specifically. Median survival was around 11 years for all patients. For CRCS, median survival is not defined since more than 50% of patients had not died due to CRC at the end of the period.

**Table 3:** Survival Outcomes

| Total cohort N = 26,204 |  |  |  |  |  |  |
| --- | --- | --- | --- | --- | --- | --- |
|  | All deaths |  |  | CRC deaths |  |  |
|  | All | Colon | Rectal | All | Colon | Rectal |
| **Number of patients** | 26,204 | 19,284 | 6,920 | 26,204 | 19,284 | 6,920 |
| **Number of deaths** | 9,094 | 6,930 | 2,164 | 5,040 | 3,722 | 1,318 |
| **Median survival** | 11.10 | 10.71 | 12.36 | - | - | 0- |
| **Median follow up** | 1,051 | 1,021 | 1,133 | 1,051 | 1,051 | 1,133 |
| **3 year survival** | 82% | 81% | 85% | 87% | 87% | 90% |
| **5 year survival** | 72% | 71% | 75% | 81% | 81% | 82% |
| **10 year survival** | 54% | 53% | 57% | 74% | 74% | 73% |

Fig. 2 presents the Kaplan-Meier (KM) survival curves for OS and CRC. 5-year OS for patients diagnosed with colon cancer was 71% and 75% for patients with a rectal cancer diagnosis. This was 81% and 82% respectively for CRCS. Fig. 3 and Fig. 4 plot the KM survival curves, again for OS and CRCS for both types of CRC, by stage at diagnosis. Survival outcomes worsen in line with the stage of disease at diagnosis and the differences between the survival functions are statistically significant.

**Figure 2:**
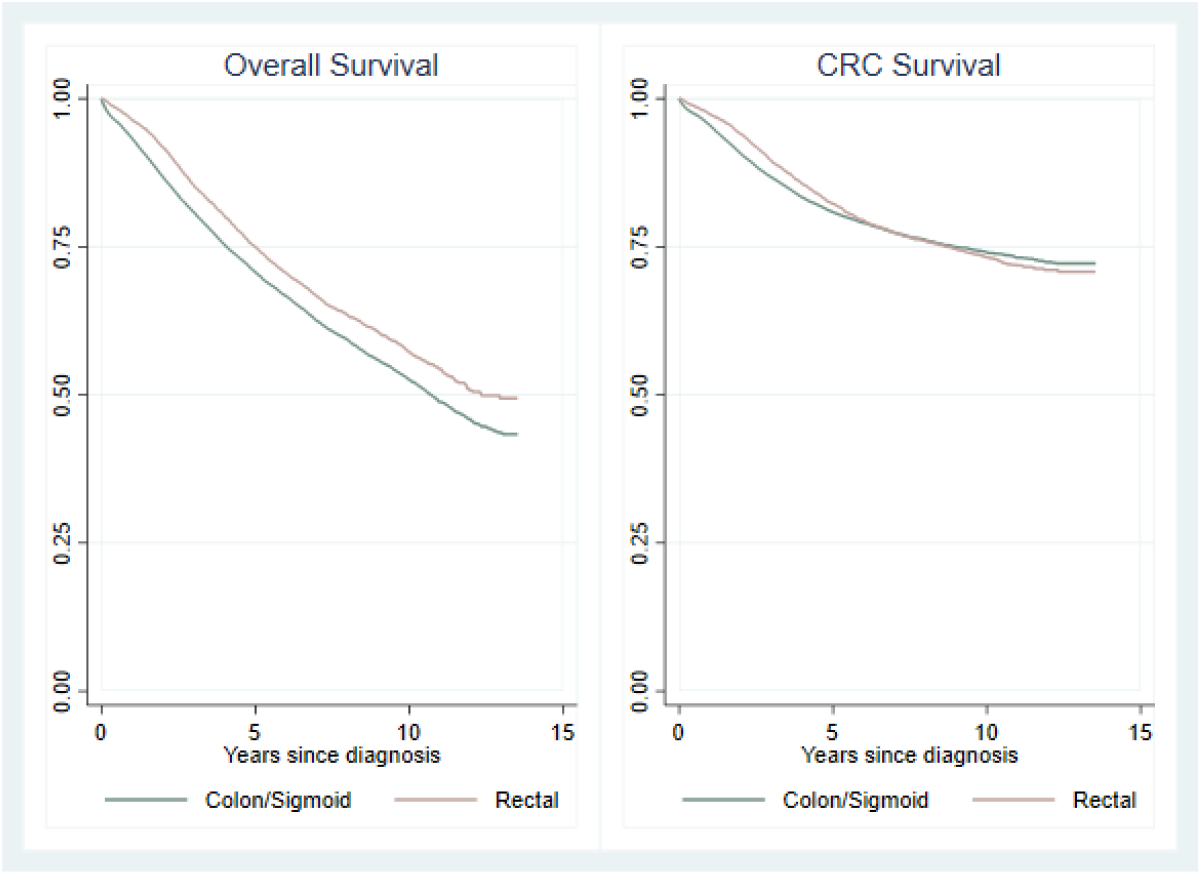
Kaplan-Meier Survival Curves by CRC types

**Figure 3:**
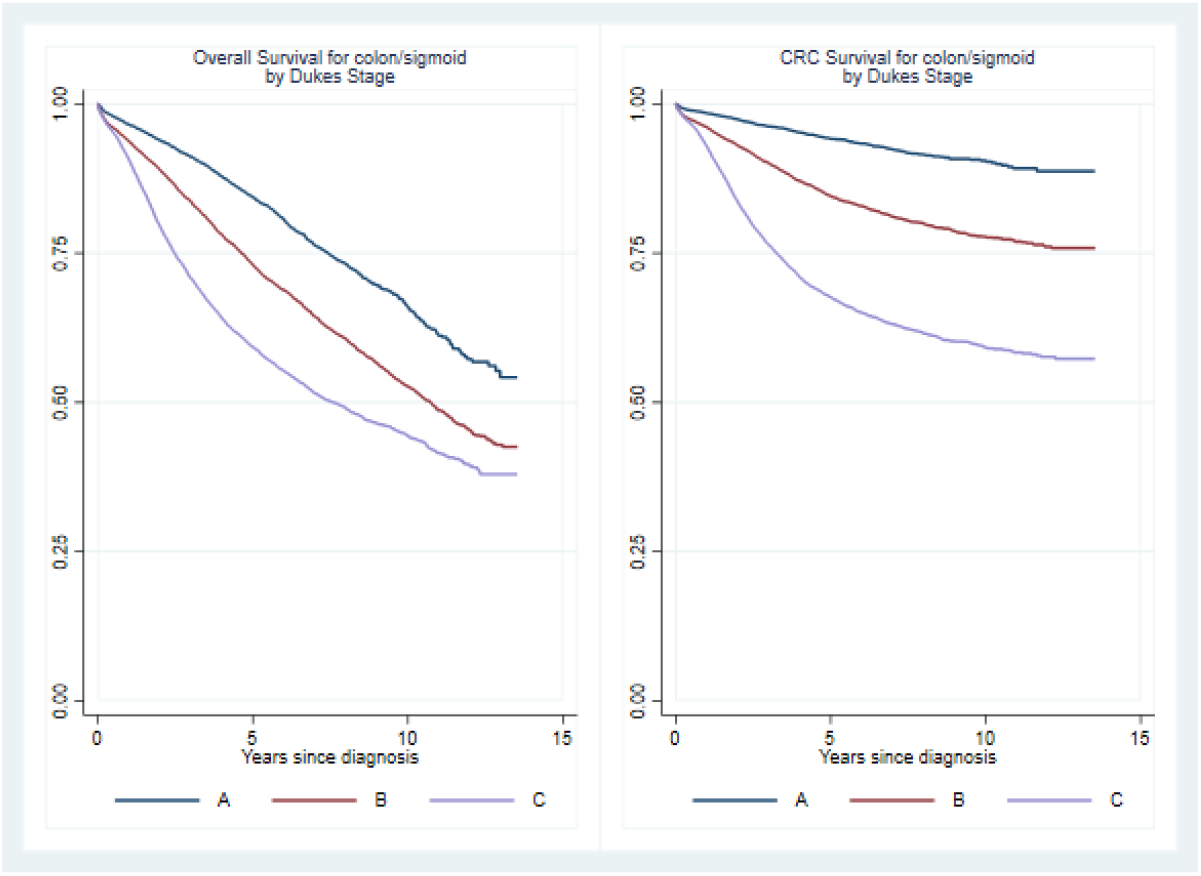
Kaplan-Meier Survival Curves for Colon patients by Dukes’s Stage

**Figure 4:**
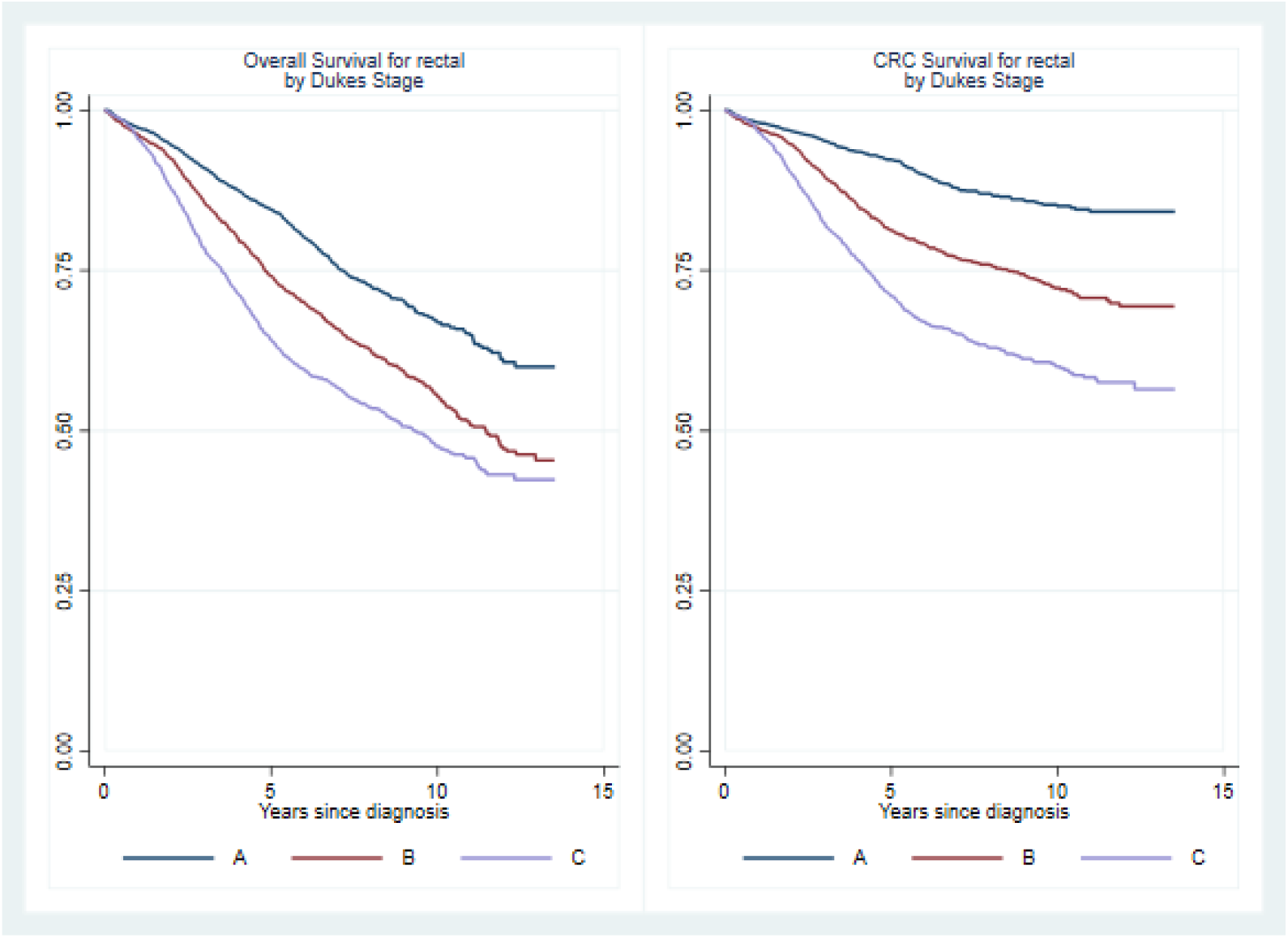
Kaplan-Meier Survival Curves for Rectal patients by Dukes’s Stage

Table 5 displays the Cox-Proportional Hazard results for patients diagnosed with colon cancer. Adjusted OS and CRCS shows that females have a significantly better survival compared to males (OS HR: 0.847, 95% CI: 0.808-0.889 and CRCS HR: 0.891, 95% CI: 0.834-0.951). Compared to those aged between 18 and 34, those aged 75 and above have a significantly higher likelihood of death and this risk increases with age. The adjusted OS HR for those aged 75-84 is 4.294 (95% CI: 2.690-6.853) and for CRCS it is 2.438 (95% CI: 1.474-4.031). The HRs for both OS and CRCS are even higher for those aged 85 and above.

**Table 4:** Cox-Proportional Hazard Regressions: Patients with colon cancer, n = 19,294

| Variable | Category | Overall Survival (OS) |  |  |  |  |  | CRC Survival (CRCS) |  |  |  |  |  |
| --- | --- | --- | --- | --- | --- | --- | --- | --- | --- | --- | --- | --- | --- |
|  |  | Unadjusted |  |  | Adjusted |  |  | Unadjusted |  |  | Adjusted |  |  |
|  |  | HR | 95 % CI |  | HR | 95 % CI |  | HR | 95 % CI |  | HR | 95 % CI |  |
| Sex | Male | 1 | - | - | 1 | - | - | 1 | - | - | 1 | - | - |
|  | Female | **0.929** | **0.886** | **0.974** | **0.847** | **0.808** | **0.889** | 0.949 | 0.889 | 1.012 | **0.891** | **0.834** | **0.951** |
| Age | 18-34 | 1 | - | - | 1 | - | - | 1 | - | - | 1 | - | - |
|  | 35-44 | 1.119 | 0.655 | 1.911 | 0.949 | 0.553 | 1.627 | 1.187 | 0.695 | 2.026 | 0.930 | 0.521 | 1.658 |
|  | 45-54 | 1.215 | 0.753 | 1.961 | 1.105 | 0.682 | 1.788 | 1.258 | 0.779 | 2.029 | 1.074 | 0.641 | 1.801 |
|  | 55-64 | 1.540 | 0.965 | 2.458 | 1.434 | 0.895 | 2.298 | 1.350 | 0.846 | 2.155 | 1.222 | 0.736 | 2.029 |
|  | 65-74 | **2.219** | **1.394** | **3.530** | **2.132** | **1.335** | **3.404** | 1.509 | 0.948 | 2.401 | 1.440 | 0.871 | 2.382 |
|  | 75-84 | **4.492** | **2.825** | **7.143** | **4.294** | **2.690** | **6.853** | **2.580** | **1.622** | **4.102** | **2.438** | **1.474** | **4.031** |
|  | 85+ | **7.914** | **4.964** | **12.617** | **7.721** | **4.823** | **12.361** | **3.889** | **2.440** | **6.201** | **3.737** | **2.244** | **6.225** |
| Cancer Network | SCAN | 1 | - | - | 1 | - | - | 1 | - | - | 1 | - | - |
|  | WOSCAN | **0.924** | **0.875** | **0.976** | 0.952 | 0.900 | 1.007 | **0.878** | **0.814** | **0.946** | **0.906** | **0.839** | **0.979** |
|  | NOSCAN | 0.987 | 0.929 | 1.049 | 1.010 | 0.949 | 1.075 | 0.989 | 0.912 | 1.073 | 0.998 | 0.917 | 1.086 |
| SIMD | 1 | 1 | - | - | 1 | - | - | 1 | - | - | 1 | - | - |
|  | 2 | **0.915** | **0.851** | **0.983** | **0.889** | **0.826** | **0.958** | 0.958 | 0.868 | 1.057 | 0.924 | 0.835 | 1.023 |
|  | 3 | **0.838** | **0.779** | **0.901** | **0.811** | **0.750** | **0.878** | **0.887** | **0.803** | **0.979** | **0.839** | **0.753** | **0.934** |
|  | 4 | **0.731** | **0.677** | **0.789** | **0.727** | **0.670** | **0.790** | **0.750** | **0.675** | **0.833** | **0.725** | **0.647** | **0.812** |
|  | 5 | **0.704** | **0.652** | **0.761** | **0.675** | **0.623** | **0.731** | **0.765** | **0.689** | **0.850** | **0.735** | **0.659** | **0.819** |
| Dukes Stage | A | 1 | - | - | 1 | - | - | 1 | - | - | 1 | - | - |
|  | B | **1.631** | **1.518** | **1.751** | **1.420** | **1.325** | **1.522** | **2.586** | **2.271** | **2.945** | **2.327** | **2.042** | **2.650** |
|  | C | **2.409** | **2.241** | **2.590** | **2.385** | **2.222** | **2.560** | **5.765** | **5.080** | **6.542** | **5.661** | **4.988** | **6.424** |
| Urban/Rural | Urban | 1 | - | - | 1 | - | - | 1 | - | - | 1 | - | - |
|  | Remote/rural | **0.930** | **0.879** | **0.984** | 0.954 | 0.897 | 1.016 | 0.955 | 0.885 | 1.030 | 0.990 | 0.909 | 1.078 |
| Year of diagnosis | 2008-10 | 1 | - | - | 1 | - | - | 1 | - | - | 1 | - | - |
|  | 2009-11 | **0.843** | **0.795** | **0.894** | **0.912** | **0.859** | **0.968** | **0.874** | **0.805** | **0.948** | 0.948 | 0.873 | 1.031 |
|  | 2012-14 | **0.736** | **0.688** | **0.787** | **0.810** | **0.757** | **0.868** | **0.746** | **0.682** | **0.816** | **0.819** | **0.748** | **0.898** |
|  | 2015-18 | **0.697** | **0.640** | **0.759** | **0.750** | **0.688** | **0.819** | **0.711** | **0.639** | **0.792** | **0.761** | **0.682** | **0.848** |
| Inpatient episodes | Mean | 1.009 | 1.005 | 1.012 | 1.002 | 0.998 | 1.006 | 1.008 | 1.003 | 1.012 | 1.001 | 0.996 | 1.007 |
| Quan morbidity score | Mean | 1.401 | 0.518 | 1.939 | **1.262** | **1.171** | **1.359** | 1.227 | 1.109 | 1.357 | **1.144** | **1.022** | **1.281** |

**Table 5:** Cox-Proportional Hazard Regressions: Patients with rectal cancer, n = 6,920

| Variable | Category | Overall Survival (OS) |  |  |  |  |  | CRC Survival (CRCS) |  |  |  |  |  |
| --- | --- | --- | --- | --- | --- | --- | --- | --- | --- | --- | --- | --- | --- |
|  |  | Unadjusted |  |  | Adjusted |  |  | Unadjusted |  |  | Adjusted |  |  |
|  |  | HR | 95 % CI |  | HR | 95 % CI |  | HR | 95 % CI |  | HR | 95 % CI |  |
| Sex | Male | 1 | - | - | 1 | - | - | 1 | - | - | 1 | - | - |
|  | Female | **0.807** | **0.739** | **0.883** | **0.761** | **0.695** | **0.834** | **0.792** | **0.706** | **0.889** | **0.744** | **0.662** | **0.838** |
| Age | 18-34 | 1 | - | - | 1 | - | - | 1 | - | - | 1 | - | - |
|  | 35-44 | 1.525 | 0.713 | 3.263 | 1.516 | 0.700 | 3.281 | 1.229 | 0.580 | 2.605 | 1.234 | 0.568 | 2.678 |
|  | 45-54 | 1.024 | 0.496 | 2.114 | 0.974 | 0.465 | 2.040 | 0.784 | 0.385 | 1.597 | 0.758 | 0.361 | 1.591 |
|  | 55-64 | 1.279 | 0.627 | 2.607 | 1.219 | 0.589 | 2.523 | 0.932 | 0.465 | 1.871 | 0.909 | 0.439 | 1.880 |
|  | 65-74 | 1.762 | 0.867 | 3.583 | 1.717 | 0.832 | 3.544 | 0.961 | 0.480 | 1.924 | 0.977 | 0.473 | 2.019 |
|  | 75-84 | **3.289** | **1.618** | **6.686** | **3.268** | **1.583** | **6.749** | 1.691 | 0.844 | 3.389 | 1.765 | 0.854 | 3.649 |
|  | 85+ | **6.520** | **3.170** | **13.410** | **7.028** | **3.362** | **14.691** | **3.375** | **1.646** | **6.919** | **3.836** | **1.811** | **8.126** |
| Cancer Network | SCAN | 1 | - | - | 1 | - | - | 1 | - | - | 1 | - | - |
|  | WOSCAN | 0.958 | 0.867 | 1.059 | 0.997 | 0.899 | 1.106 | 0.935 | 0.823 | 1.064 | 0.974 | 0.852 | 1.113 |
|  | NOSCAN | 1.073 | 0.966 | 1.192 | **1.138** | **1.020** | **1.271** | 1.117 | 0.977 | 1.277 | **1.180** | **1.025** | **1.357** |
| SIMD | 1 | 1 | - | - | 1 | - | - | 1 | - | - | 1 | - | - |
|  | 2 | **0.875** | **0.768** | **0.997** | 0.862 | 0.754 | 0.987 | 0.882 | 0.743 | 1.046 | 0.879 | 0.737 | 1.048 |
|  | 3 | 0.888 | 0.780 | 1.012 | 0.925 | 0.805 | 1.064 | 0.919 | 0.776 | 1.089 | 0.952 | 0.796 | 1.139 |
|  | 4 | **0.794** | **0.696** | **0.906** | **0.795** | **0.689** | **0.918** | 0.854 | 0.720 | 1.012 | **0.832** | **0.694** | **0.999** |
|  | 5 | **0.743** | **0.647** | **0.853** | **0.737** | **0.639** | **0.852** | **0.794** | **0.664** | **0.948** | **0.789** | **0.656** | **0.949** |
| Dukes Stage | 1 | 1 | - | - | 1 | - | - | 1 | - | - | 1 | - | - |
|  | 2 | **1.542** | **1.385** | **1.718** | **1.456** | **1.308** | **1.621** | **2.099** | **1.799** | **2.449** | **1.993** | **1.707** | **2.327** |
|  | 3 | **2.060** | **1.854** | **2.290** | **2.223** | **2.001** | **2.470** | **3.440** | **2.975** | **3.977** | **3.594** | **3.107** | **4.157** |
| Urban/Rural | Urban | 1 | - | - | 1 | - | - | 1 | - | - | 1 | - | - |
|  | Remote/rural | 0.984 | 0.893 | 1.084 | 0.943 | 0.846 | 1.053 | 1.026 | 0.907 | 1.161 | 0.962 | 0.836 | 1.108 |
| Year of diagnosis | 2008-10 | 1 | - | - | 1 | - | - | 1 | - | - | 1 | - | - |
|  | 2009-11 | 0.950 | 0.856 | 1.054 | 0.987 | 0.888 | 1.097 | 0.889 | 0.777 | 1.017 | 0.920 | 0.802 | 1.054 |
|  | 2012-14 | **0.800** | **0.708** | **0.905** | **0.866** | **0.764** | **0.982** | **0.785** | **0.674** | **0.913** | **0.854** | **0.732** | **0.997** |
|  | 2015-18 | **0.674** | **0.570** | **0.798** | **0.760** | **0.640** | **0.901** | **0.732** | **0.601** | **0.891** | **0.812** | **0.666** | **0.990** |
| Inpatient episodes | Mean | **1.007** | **1.004** | **1.010** | 1.003 | 0.999 | 1.007 | **1.007** | **1.003** | **1.012** | 1.002 | 0.997 | 1.008 |
| Quan morbidity score | Mean | **1.477** | **1.315** | **1.659** | **1.317** | **1.159** | **1.495** | **1.361** | **1.157** | **1.602** | **1.252** | **1.049** | **1.495** |
| Significance at the 5% level denoted by bold text |  |  |  |  |  |  |  |  |  |  |  |  |  |

In terms of geography, by cancer network, WoSCAN patients have significantly reduced risk of death from CRC relative to patients in SCAN and this difference remains significant after adjusting for other factors (adjusted HR: 0.906, 95% CI: 0.839-0.979). The adjusted models show that living in a remote or rural area has no influence on risk of death.

The adjusted OS results show that the risk of dying decreases with lower levels of deprivation. Compared to the most deprived quintile, the HR for those living in the second most deprived quintile is 0.889 (95% CI: 0.826-0.958), whilst the HR for those living in the least deprived quintile is 0.675 (95% CI: 0.623-0.731). A similar trend was observed for CRCS, with the adjusted model showing a lower risk of death with reduced deprivation, however no difference in risk of death was observed between the first and second most deprived quintiles.

All models demonstrate an increased risk of death with higher disease stage. In the adjusted models, compared to those with stage I disease, the HR’s for risk of death from any cause for those with stage II or III disease are 1.42 (95% CI: 1.325-1.522) and 2.385 (95% CI: 2.222-2.560) respectively, and for CRC specific deaths, the HR’s are 2.327 (95% CI: 2.042-2.650) and 5.661 (95% CI: 4.988-6.424) respectively.

The year of diagnosis also has a significant influence on the risk of death. The adjusted HRs show a significant reduction in risk of death over time and thus an improvement in survival. For example, the risk of death from CRC was lower if a patient was diagnosed in 2015-18 compared to if they were diagnosed in 2006-08 (HR: 0.761, 95% CI: 0.682-0.848).

In terms of inpatient episodes within the five years pre-diagnosis, all models show no significant increase in the likelihood of death as the number of hospital episodes increases.

As for comorbidity, the models for OS show an increased risk of death as co-morbidity increases, even after accounting for other factors. Specifically, the HR on the Quan co-morbidity score variable is 1.262 (95% CI: 1.171-1.359). Similarly, in the models for CRCS, the risk of death increases as co-morbidity increases (HR: 1.144, 95% CI: 1.022-1.281).

Table 5 displays the Cox-Proportional Hazard results for patients diagnosed with rectal cancer. The results from the adjusted models for OS and CRC show that females have a lower risk of death compared to males (OS HR: 0.761, 95% CI: 0.695-0.834; CRCS HR: 0.744, 95% CI: 0.662-0.838). After controlling for other factors, older age groups have a higher risk of death from both all causes and CRC specifically. For those aged 85 and over, the adjusted model HR’s for OS and CRCS are 7.028 (95% CI: 3.362-14.691) and 3.836 (95% CI: 1.811-8.126) respectively.

The adjusted models for patients diagnosed with rectal cancer suggest that compared to patients diagnosed in SCAN, there is no significant difference in risk of death compared to patients diagnosed in WOSCAN. However, rectal cancer patients diagnosed in NOSCAN have a higher risk of death. The HRs are 1.138 (95% CI: 1.020-1.271) and 1.180 (95% CI: 1.025-1.357) for OS and CRCS respectively.

Upon adjusting for other factors, the OS model suggests that the risk of death from any cause is significantly lower for the two least deprived areas (HR for quintile 4: 0.795, 95% CI: 0.689-0.918, HR for qunitile 5: 0.737, 95% CI:0.639-0.852). Similarly, a significant difference is observed for CRCS between the most and the two least deprived quintiles (HR for quintile 4: 0.832, 95% CI: 0.694-0.999, HR for quintile 5: 0.789, 95% CI: 0.656-0.949).

Stage of disease at diagnosis is associated with an increasing risk of death. The HRs in the adjusted models for stage II and III are 1.456 (95% CI: 1.308-1.621) and 2.223 (95% CI: 2.001-2.470) respectively for OS. For CRCS, they are 1.993 (95% CI: 1.707-2.327) and 3.594 (95% CI: 3.107-4.157), again for stage II and III respectively.

Survival outcomes also improved over time, with diagnoses in later years associated with better outcomes. In particular, compared to those diagnosed in 2008-10, those diagnosed in 2015-18 had significantly better OS and CRCS (OS HR: 0.760, CI: 0.640-0.901; CRCS HR: 0.812, CI: 0.666-0.990).

In the five years pre-diagnosis, an increase in the number of hospital inpatient episodes had no effect on risk of death from any cause or CRC, once other factors were controlled for. Quan co-morbidity score however, was associated with a significant increase in the risk of death (OS HR: 1.317, CI: 1.159-1.495; CRCS HR: 1.252, CI: 1.049-1.495).

## 4. DISCUSSION

This paper used a national linked administrative dataset to provide up to date, real world evidence on the treatment and survival outcomes for patients diagnosed with CRC. We described the demographic, diagnostic and treatment characteristics of patients with colon cancer and rectal cancer. Further, for those patients treated on a curative pathway, we assessed the factors affecting patients overall and CRC specific survival.

Cox-proportional hazard models for a curative cohort of patients with colon or rectal cancer confirm that, older individuals are at increased risk of death. Further, the stage at diagnosis influences survival, with those diagnosed at later stages having poorer survival compared to those diagnosed at the earliest stage. The adjusted models for OS show that patients diagnosed with stage III colon or rectal cancer have a twofold increase in risk of death when compared to those diagnosed at stage I. For CRC specific deaths, this risk increases to almost sixfold for patients diagnosed with colon cancer and almost fourfold for those diagnosed with rectal cancer.

The models show that in general, survival has improved consistently since 2006-08 for patients diagnosed with either colon or rectal cancer. Furthermore, as expected, the models find a higher co-morbidity score is associated with significantly poorer survival outcomes for patients diagnosed with both colon and rectal cancer.

Despite controlling for underlying patient co-morbidity, the models also show a worrying result in that they suggest significant regional variation in outcomes between the three MCNs. In particular, patients diagnosed with colon cancer in WoSCAN appear to have better CRC survival outcomes compared to patients in SCAN. At the same time, patients diagnosed with rectal cancer in NoSCAN appear to have significantly poorer survival outcomes, both all cause and CRC specifically, compared to those in SCAN. These differences suggest potentially inequitable outcomes for patients in an NHS that is designed to ensure equal access to quality care. This finding requires further investigation to understand why we see significant variation in survival between the MCNs, despite controlling for disease stage and patient needs. The models also account for location via the urban/rural indicator. In contrast to previous literature, the models do not find any evidence that rurality has a significant impact on survival (Campbell et al., 2000).

Another concerning finding from our study is that the level of deprivation in the local area is significantly associated with poorer survival outcomes, even after accounting for clinical factors and patient need. This finding is consistent with some previous research carried out in Scotland (Shack et al., 2007). The reasons for poorer outcomes in the most deprived areas in Scotland is potentially due to later presentation or delay in treatment, however recent evidence from one MCN in Scotland finds that there is no association between deprivation and these factors (Paterson et al., 2014). Another possible explanation for the observed differences could be other co-morbidities present within more deprived populations, though we attempt to control for this effect via measures from hospital admissions data. Once again, further investigation into these differences is warranted. In particular, an investigation into type, length and timing of treatment. In addition, the models find evidence that women have a significant survival advantage compared to men. This result is consistent with previous evidence from Scotland and around the world (McArdle et al., 2003; Yang et al., 2017). The explanation for these differences remains unclear but might be explained partly by endogenous factors such as genetics or hormones, particularly those present in younger females (Majek et al., 2013).

In summary, we have used a nationally linked administrative data set to retrospectively explore patterns in treatment and outcomes for CRC patients in Scotland. We have demonstrated that Scotland’s unique data linkage infrastructure can accommodate linkage between demographic records, cancer registry and hospital admissions data, providing a fuller picture of the needs of CRC patients. We have identified a number of areas which require further research including regional and socio-demographic differences in outcomes.

Key next steps in our analysis will be to investigate these areas further and utilise a number of additional administrative data sets that have been linked to the registry data for the purposes of this project (Hanna et al., 2021). These include detailed information on chemotherapy prescribing and cancer audit data.

## Data Availability

The data used in this project are not available for re-use.

